# Machine learning for classifying chronic kidney disease and predicting creatinine levels using at-home measurements

**DOI:** 10.1101/2024.03.15.24304364

**Authors:** Brady Metherall, Anna K. Berryman, Georgia S. Brennan

## Abstract

**Background:** Chronic kidney disease (CKD) is a global health concern with early detection playing a pivotal role in effective management. Machine learning models demonstrate promise in CKD detection, yet the impact on detection and classification using different sets of clinical features remains under-explored.

**Methods:** In this study, we focus on CKD classification and creatinine prediction using three sets of features; at-home, monitoring, and laboratory. We employ artificial neural networks (ANNs) and random forests (RFs) on a dataset of 400 patients with 25 input features, which we divide into three feature sets. Using 10-fold cross-validation, we calculate metrics such as accuracy, true positive rate (TPR), true negative rate (TNR), and mean squared error.

**Results:** Our results reveal RF achieves superior accuracy (92.5%) in at-home CKD classification over ANNs (82.9%). ANNs achieve a higher TPR (92.0%) but a lower TNR (67.9%) compared with RFs (90.0% and 95.8%, respectively). For monitoring and laboratory features, both methods achieve accuracies exceeding 98%. The R2 score for creatinine regression is approximately 0.3 higher with laboratory features than at-home features. Feature importance analysis identifies key clinical variables hemoglobin and blood urea, and key comorbidities hypertension and diabetes mellitus, in agreement with previous studies.

**Conclusions:** Machine learning models, particularly RFs, exhibit promise in CKD diagnosis and highlight significant features in CKD detection. Moreover, such models may assist in screening a general population using at-home features—potentially increasing early detection of CKD, thus improving patient care and offering hope for a more effective approach to managing this prevalent health condition.

## 1 Introduction

Chronic kidney disease (CKD) represents a global health challenge affecting millions worldwide and placing a substantial burden on healthcare systems [1, 2]. More women are affected by CKD than breast cancer, and more men than prostate cancer [3]. CKD is often a silent and progressive condition remaining undetected until a significant loss of kidney function has occurred. Early detection and prediction are crucial for timely interventions and improved patient outcomes.

CKD is classified into five stages [4–6] based on glomerular filtration rate (GFR)—a measure of kidney function. GFR measurement is complex and so is usually estimated using equations. The two most common equations are the MDRD [7] and CKD-EPI [8] equations. The MDRD equation has limitations for healthy individuals or those with mild kidney dysfunction. In contrast, the CKD-EPI equation addresses some of these limitations, providing a more accurate estimate of GFR by adjusting for different creatinine ranges.

Machine learning, which has shown success in predicting diseases [9], holds promise within nephrology [10] including enhancing CKD screening and detection [3, 11–13]. One promising application of machine learning in the context of CKD is the potential for at-home detection or screening. At-home CKD screening offers an ideal solution to the global health challenge posed by CKD. By leveraging user-friendly devices and predictive models, individuals could track key indicators of kidney health in the comfort of their homes. This approach not only increases early detection but also enables a wider subset of the population to be screened. The convenience of at-home screening, coupled with the predictive capabilities of machine learning, could significantly improve the efficiency of CKD management, leading to better patient outcomes and alleviating the burden on healthcare systems worldwide. Moreover, such an approach aligns with the broader trend of personalized and preventive healthcare.

Our primary focus is detection of CKD with at-home measurements to facilitate easier and earlier detection throughout the general population. Online CKD detection, such as through a health application on smartphones, is identified as an area for future research by Qezelbash-Cham *et al*. [3]. However, such an application would be limited to more accessible features than in a clinical setting [11]. To the best of our knowledge there is no study comparing the performance of machine learning algorithms using the full feature set and only the features that can be measured at home. To gain insight into the possible utility of machine learning for at-home detection we develop machine learning models for two purposes. First, we aim to classify whether a patient has CKD (by grouping all five stages together), and second, to predict creatinine levels, as is done in [14], which can then be used to calculate the estimated GFR and consequently determine the stage of CKD. In this study, we categorize the features into three sets: at-home features (tests that can be easily conducted at home), monitoring features (encompassing basic testing), and laboratory features (including comprehensive tests). To achieve these goals we employ artificial neural networks and random forest. Artificial neural networks and random forest are two of the most commonly investigated and successful machine learning algorithms in the context of chronic kidney disease according to the review [12].

## 2 Methods

### 2.1 Data

We use a publicly available dataset hosted on the UCI machine learning repository [15]. The dataset consists of 400 patients from a hospital in Tamil Nadu, India admitted over a period of two months. The dataset has been widely used to apply machine learning techniques to CKD detection and classification; over 50% of the studies in the review [3] used this dataset. The data on each patient includes blood test results, in addition to comorbidity and demographic data. The data has 250 patients labelled as having CKD and 150 patients labelled without CKD. The labels were assigned by nephrologists based on patient history, symptoms, and blood and urine tests [16]. The data also includes serum creatinine, which is the key quantity in both the MDRD and CKD-EPI equations for estimated GFR. Furthermore, we supplement the data with sex and race data from [17]. We will use both the CKD indicator and serum creatinine measurement as dependent variables in our analysis. For descriptive statistics of the data see Appendix A.

#### 2.1.1 Feature sets

To help facilitate at-home diagnoses we separate the features into three sets as shown in Table 1. The smallest set is made up of features measurable at home (at-home features). At-home features include all patient demographic and comorbidity data, as well as blood pressure, which can be easily measured either at home or at a local pharmacy. The second set, monitoring features, are typically obtained if a patient is monitoring their health with health checks in a clinic. The monitoring set includes the at-home features as well as features measurable with standard tests (*i*.*e*. blood urea and blood glucose). Finally, the third set, which we call laboratory features, includes all 25 features, where the remaining features are measured with more specialized tests.

**Table 1:** Feature list organized into at-home, monitoring, and laboratory groups, where the at-home group is a subgroup of monitoring, and monitoring is a subgroup of laboratory features.

| Attribute | Symbol | Type | Feature set |
| --- | --- | --- | --- |
| Age | age | numerical | at-home |
| Race | race | nominal | at-home |
| Sex | sex | nominal | at-home |
| Hypertension | htn | nominal | at-home |
| Diabetes mellitus | dm | nominal | at-home |
| Coronary artery disease | cad | nominal | at-home |
| Appetite | appet | nominal | at-home |
| Pedal edema | pe | nominal | at-home |
| Anemia | ane | nominal | at-home |
| Blood pressure | bp | numerical | at-home |
| Red blood cells | rbc | nominal | monitoring |
| Red blood cell count | rbcc | numerical | monitoring |
| White blood cell count | wbcc | numerical | monitoring |
| Blood glucose random | bgr | numerical | monitoring |
| Blood urea | bu | numerical | monitoring |
| Sodium | sod | numerical | monitoring |
| Potassium | pot | numerical | monitoring |
| Hemoglobin | hemo | numerical | monitoring |
| Specific gravity | sg | nominal | laboratory |
| Albumin | al | nominal | laboratory |
| Sugar | su | nominal | laboratory |
| Bacteria | ba | nominal | laboratory |
| Pus cell | pc | nominal | laboratory |
| Pus cell clumps | pcc | nominal | laboratory |
| Packed cell volume | pcv | numerical | laboratory |

#### 2.1.2 Pre-processing

Our pre-processing of the data involves three steps. First, we discard the 17 patients that do not have a serum creatinine reading since we cannot use these patients to train or test our regression models. Second, we impute the missing values of numerical features. Qin *et al*. [18] investigated the impact of missing data imputation techniques on the prediction of CKD. They suggest using *k*-nearest neighbours (*k*-NN) imputation instead of mean imputation because of potential skewness in the data. The *k*-nearest neighbours algorithm finds the *k*-nearest neighbours in the space of complete features. The mean of these *k* neighbours is then used to impute the missing data. We use *k*-NN imputing with *k* = 5 to deal with missing data in numerical features. Finally, we use one hot encoding to represent nominal features. One hot encoding takes nominal features with *n* classes and replaces the original feature column in the feature matrix with *n* columns each representing a single class. Each row contains one 1 in the column that corresponds to a patient’s class, and 0s in all the remaining columns. We treat any missing values in the nominal features as their own class in the one hot encoding instead of imputing their value.

We standardize the numerical features to have mean 0 and standard deviation 1 so the distribution of values is comparable across features. Furthermore, we use the log of serum creatinine for the regression. This ensures we predict only positive values for serum creatinine and gives us more uniformly distributed data.

### 2.2 Machine learning algorithms

To evaluate the potential for at-home, early detection of CKD, we undertake two investigations: first, focusing on the classification of CKD, and second, directly predicting creatinine levels. We investigate these two tasks on each feature set described in Table 1. To address these challenges, we employ two machine learning techniques: artificial neural networks (ANNs) and random forests (RFs).

ANNs are promising tools in clinical medicine and are inspired by biological neural networks. They consist of interconnected nodes organized into layers to process data and learn patterns. ANNs excel at recognizing complex patterns, making accurate predictions, and adapting to change. ANNs’ architecture includes input, hidden, and output layers, with weighted connections adjusted during training. However, challenges exist; ANNs often require large amounts of high-quality, diverse training data for optimal performance and generalization. They can lack interpretability, acting as black boxes, making decision processes unclear.

Alongside ANNs, we will test RF algorithms, an ensemble method combining multiple decision trees trained on subsets of the data and features. During prediction, RFs aggregate tree predictions using majority voting or averaging, thus reducing overfitting and enhancing generalization. RFs generally provide better interpretability than ANNs. The decision trees in RFs can be visualized, revealing learned rules and conditions, and measures of feature importance indicate each feature’s contribution.

### 2.3 Model evaluation

We now introduce the loss functions we employ to optimize our models with respect to, as well as the metrics we will use to assess our model’s ability to classify CKD and predict creatinine levels. For binary classification, we use the loss function of cross-entropy, or log loss. Within an ANN, entropy gauges the disparity between the predicted probability of a binary outcome and the actual binary label, and for RF entropy determines the splitting criteria within a decision tree. In our evaluation of classification models on the test data we will employ five essential metrics. The primary metric is accuracy, which provides a basic measure of overall classification correctness but may fall short in imbalanced class scenarios. Additionally, we use the true positive rate (TPR) to gauge the model’s effectiveness in correctly identifying positive cases and the true negative rate (TNR) to assess its proficiency in recognizing negative cases. We also consider the false positive rate (FPR) to measure the proportion of incorrect positive classifications. Finally, the false negative rate (FNR) to evaluate the model’s ability to avoid missing positive cases, especially where false negatives could have significant consequences, such as our application. These metrics collectively offer a comprehensive evaluation of a classification models’ performance. Receiver operating characteristics (ROC) curves are commonly used in medical decision making and increasingly in machine learning since simple accuracy is often a poor metric [19]. Typical machine learning classification algorithms yield a probability of a sample being in any class. Thus, the discrete class assigned to each sample depends on the threshold used for assigning a discrete class from the probability. ROC curves plot the true positive rate against the false positive rate as the threshold varies. ROC curves allow us to understand the ability of a classifier to rank positive instances above negative instances [19]. We follow the suggestion in [20] and use vertical averaging to find the mean ROC curve across folds. It is common to reduce a ROC curve down to a single number—the area under the curve (AUC). The AUC is always between 0 and 1, where 1 is perfect performance, 0.5 is random guessing, and 0 *always mis-classifies* (which can be inverted to yield perfect performance).

To evaluate the performance of our creatinine regression models we employ three commonly used metrics. Mean squared error (MSE) measures the average squared difference between predicted and true values and has a sensitivity to larger errors and outliers. The R-squared (R2) score indicates the proportion of variance explained by the model—a higher score signifies a better fit to the data. Finally, mean absolute error (MAE) computes the average absolute difference between predicted and true values, offering robustness against outliers. In our creatinine regression experiments, we optimize our models by minimizing MSE, ensuring the models provide accurate predictions while being sensitive to potential outliers in the dataset.

#### 2.3.1 *k*-fold cross-validation

Owing to our small dataset, we use *k*-fold cross-validation, a method that involves splitting the data into *k* subsets, or folds, for iterative model training and evaluation. *k*-fold cross-validation ensures better data utilization, and reduces over-fitting risks associated with small datasets by using each sample for training and validation across different iterations. This approach enhances the reliability of performance estimation. Additionally, *k*-fold cross-validation fosters robustness in performance evaluation, overcoming sample-dependency issues present in single train/test splits. Averaging performance metrics from the *k* folds provides a more stable probabilistic performance assessment. Furthermore, *k*-fold cross-validation aids in model selection and hyperparameter tuning when working with limited data, enabling fair comparisons and effective optimization.

### 2.4 Model training

We first randomly split our data into 10 folds to use *k*-fold cross-validation with *k* = 10. In this way, we have a 90/10 train/test split for each fold. We then reserve 20% of the training data for validation. We implement our ANN models using `Keras` [21], while we use `scikit-learn` [22] for RF, both in Python. Within each fold, we tune the hyperparameters for the model. We show in Table 2 the values and ranges within the hyperparameter tuning. We use 50 trials in a random search using the `Keras` tuner [23] and `GridSearchCV` within `scikit-learn` for ANN and RF, respectively. For RFs the 50 trials are distributed among 5 inner folds. We then choose the best set of hyperparameters and re-train the model, and then evaluate the model on the test data. We repeat this process for both ANN and RF on each of the three features sets.

**Table 2:**
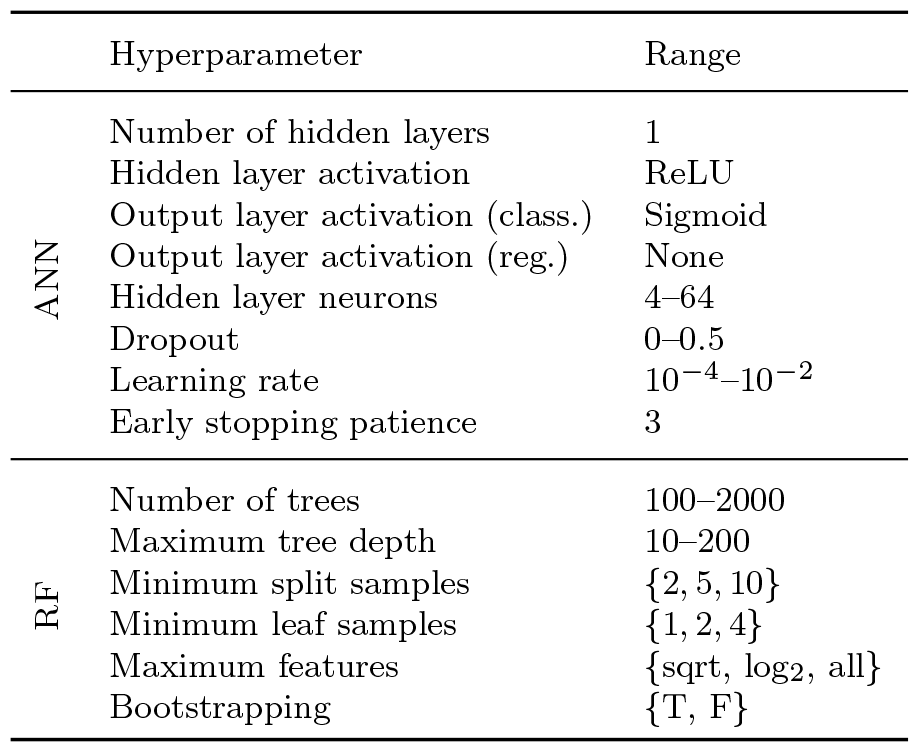
Hyperparameter values and tuning ranges within our experiments.

## 3 Results

After pre-processing the data, we are left with 383 patients with 54 features each (owing to the one hot encoding), and a split of 238/145 of CKD and not CKD. Of the 54 features, 27 and 18 are a part of the monitoring and at-home feature set, respectively. Using both ANN and RF we conduct both machine learning tasks on each feature set. We compute the metrics described in the previous section on the test data of each of the 10 folds, and compute the mean and standard deviation across the folds. We show our results using both ANN and RF to classify CKD in Table 3, and present creatinine prediction results in Table 4. Furthermore, in Figure 2, we show the feature importance extracted from the RF models.

**Table 3:** CKD binary classification metric results presented as means of the 10 folds plus–minus the standard deviation.

|  | Features | Entropy | Accuracy | TPR | TNR | FPR | FNR |
| --- | --- | --- | --- | --- | --- | --- | --- |
| ANN | at-home | $0.349 \pm 0.116$ | $82.9 \pm 9.93$ | $92.0 \pm 7.00$ | $67.9 \pm 34.8$ | $32.1 \pm 34.8$ | $8.03 \pm 7.00$ |
| | monitoring | $0.0800 \pm 0.0401$ | $98.7 \pm 2.12$ | $98.8 \pm 2.63$ | $98.5 \pm 2.95$ | $1.46 \pm 2.95$ | $1.22 \pm 2.63$ |
| | laboratory | $0.0404 \pm 0.0541$ | $99.2 \pm 1.69$ | $98.8 \pm 2.50$ | $100.0 \pm 0.00$ | $0.00 \pm 0.00$ | $1.19 \pm 2.50$ |
| RF | at-home | $0.215 \pm 0.0680$ | $92.5 \pm 4.08$ | $90.0 \pm 6.63$ | $95.8 \pm 5.50$ | $4.23 \pm 5.50$ | $9.96 \pm 6.63$ |
| | monitoring | $0.0714 \pm 0.0315$ | $98.7 \pm 1.77$ | $99.0 \pm 1.97$ | $98.0 \pm 3.08$ | $1.96 \pm 3.08$ | $0.972 \pm 1.97$ |
| | laboratory | $0.0394 \pm 0.0199$ | $99.5 \pm 1.05$ | $99.6 \pm 1.25$ | $99.2 \pm 2.50$ | $0.833 \pm 2.50$ | $0.417 \pm 1.25$ |

**Table 4:**
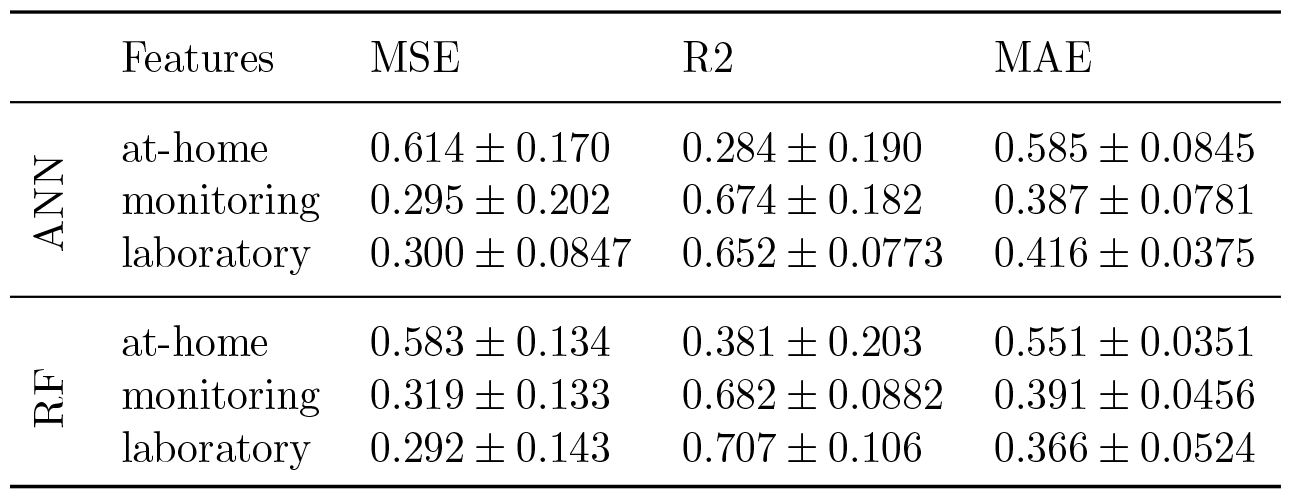
Creatinine regression metric results presented as means of the 10 folds plus–minus the standard deviation.

|  | Features | MSE | R2 | MAE |
| --- | --- | --- | --- | --- |
| ANN | at-home | $0.614 \pm 0.170$ | $0.284 \pm 0.190$ | $0.585 \pm 0.0845$ |
| | monitoring | $0.295 \pm 0.202$ | $0.674 \pm 0.182$ | $0.387 \pm 0.0781$ |
| | laboratory | $0.300 \pm 0.0847$ | $0.652 \pm 0.0773$ | $0.416 \pm 0.0375$ |
| RF | at-home | $0.583 \pm 0.134$ | $0.381 \pm 0.203$ | $0.551 \pm 0.0351$ |
| | monitoring | $0.319 \pm 0.133$ | $0.682 \pm 0.0882$ | $0.391 \pm 0.0456$ |
| | laboratory | $0.292 \pm 0.143$ | $0.707 \pm 0.106$ | $0.366 \pm 0.0524$ |

**Fig. 1:**
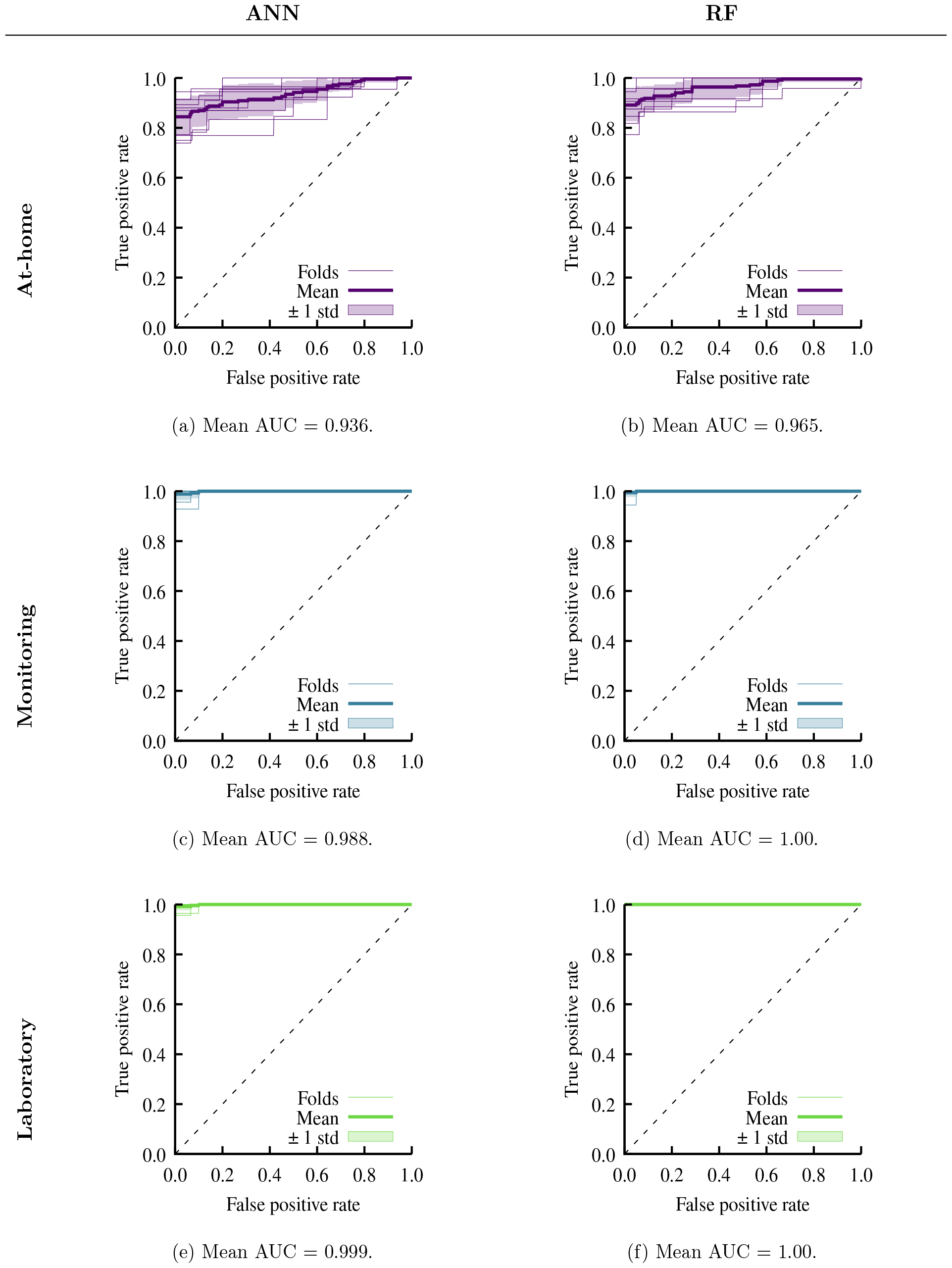
CKD binary classification ROC curves with AUC values.

**Fig. 2:**
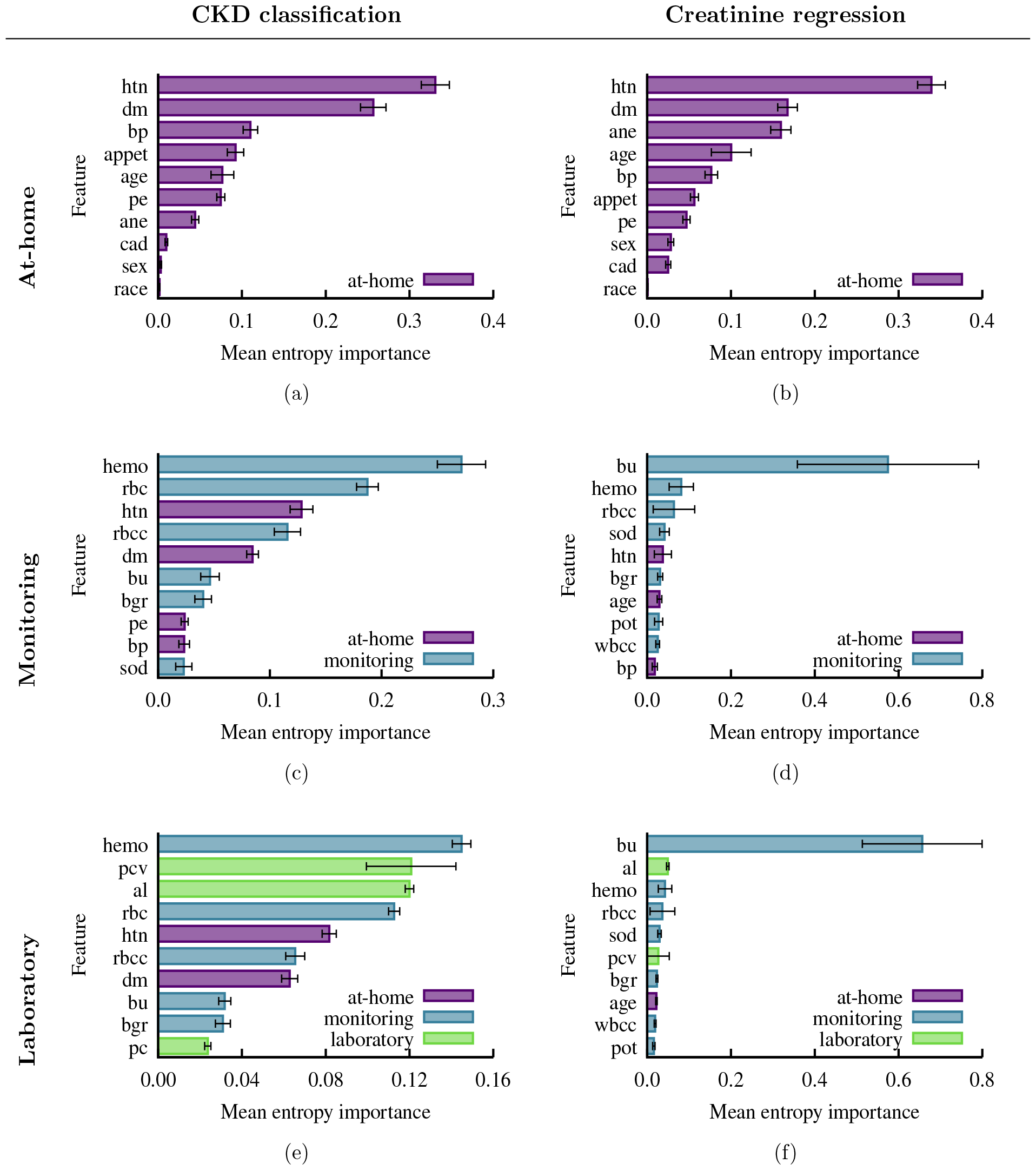
Mean entropy importance for the six experiments extracted from RF. The error bars denote the standard deviation.

### 3.1 CKD classification

We show the results of our 10-fold cross-validation CKD classification in Table 3. We observe a marked difference in CKD classification metrics between the two machine learning methods with at-home features. Using at-home features, ANN achieves an average of 82.9%, 92.0%, and 67.9% for accuracy, TPR, and TNR, respectively, over the 10 folds. On the other hand, RF recovers a higher accuracy of 92.5%, a comparable 90.0% TPR, and a significantly higher 95.8% TNR. We note that the TPR for the at-home features is higher with ANN than RF, however, the higher accuracy and TNR of RF over ANN makes RF the better algorithm for at-home CKD classification. Accuracy of both ANN and RF CKD classification for both monitoring and laboratory features is over 98%. Furthermore, the TPR and TNR for both monitoring and laboratory features is also nearly perfect with rates exceeding 98%. There is little separation in the results for these two feature sets and the two methods. RF and ANN both perform worse with at-home features than with monitoring or laboratory features, but, with the at-home feature set RF performs considerably better than ANN.

In Figure 1 we show the ROC curves from the six classification tasks. We find similar performance between ANN and RF for all three feature sets. With the at-home features RF has a slightly higher AUC of 0.965 compared to 0.936 with ANN. With monitoring or laboratory features RF has an AUC of 1.00, while ANN is 0.998 and 0.999 for monitoring and laboratory features, repsectively.

### 3.2 Creatinine regression

We now turn our attention to the task of creatinine regression; we show our results in Table 4. The ANN and RF results are similar to each other. The only noticeable difference is in the R2 score for the at-home features. Here, RF performs better, however, with an average R2 score of 0.381, creatinine levels are not well predicted. In the remaining results, the method that performs better is evenly split between ANN and RF. As we saw with the classification, we again find there are only negligible differences between the monitoring and laboratory feature metrics. Once again, we find the at-home metrics are poorer than the monitoring or laboratory results. The MSE of the at-home regression is roughly double the MSEs of the monitoring and laboratory feature sets. One interesting result is that for ANN all three metrics were better for the monitoring features than with the full laboratory features.

### 3.3 Random forest feature importance

Random forest allows us to estimate the importance of individual features from their frequency within the decision trees. In Figure 2, we show the top 10 most important features (by mean entropy importance) for the six RF experiments. In the CKD classification problem for the monitoring features (Figure 2c) and laboratory features (Figure 2e) we find hemoglobin (hemo) is the most important feature in both groups. Hemoglobin has a mean entropy importance of 0.271 and 0.145, for the monitoring and laboratory features, respectively. Out of the top 10 most important classification laboratory features only three features are exclusive to the laboratory group. The overlap with the important monitoring features may help explain the small differences in metrics between the monitoring and laboratory feature sets as we saw in Table 3. Furthermore, the normal/abnormal red blood cell feature (rbc) is the second most important feature in the monitoring set with a mean entropy importance of 0.187, as well as the fourth most important feature in the laboratory set (0.113 mean entropy importance). We note in the laboratory features the red blood cell feature is only marginally less important than the packed cell volume (pcv), which has a mean entropy importance of 0.121. The top two features in the at-home set, hypertension (htn) and diabetes mellitus (dm), also appear in the top five features in the monitoring set, and are the fifth and seventh most important features in the full laboratory set. Both hypertension and diabetes mellitus play an important role in the 92.5% accuracy achieved with RF classification.

Turning to creatinine prediction, we again find hypertension and diabetes mellitus are the two most important features in the at-home set (Figure 2b). However, anemia (ane) plays a more substantial role than in the classification task. In classification, blood urea (bu) was among the top 10 of both the monitoring and laboratory features. In the creatinine prediction however, blood urea is by far the most important feature in both sets. Moreover, hemoglobin (hemo), red blood cell count (rbcc), and sodium (sod) are each in the top 5 of important features in the monitoring and laboratory sets. Hypertension drops to fifth most important in the monitoring features, and is not within the top 10 for laboratory features.

## 4 Discussion

We separated the features into the three sets, as outlined in Table 1, based on the tools required for measurement. At-home features are known by a patient or are easily measurable, like age or blood pressure. Monitoring features are obtainable from regular check-ups, such as red blood cell count, and laboratory features are from blood and urine tests targeted towards CKD, such as urine albumin.

We have classified CKD and predicted creatinine levels on the three groups of features. In the classification case, we classified whether a person has CKD or not, disregarding stages. A patient’s GFR, and thus stage, can be estimated using the predicted creatinine level as an input into the CKD-EPI equation [8]. We carried out our experiments using both ANN and RF. Using the monitoring and laboratory feature sets, both ANN and RF had a near perfect classification accuracy, and RF performed better than ANN using the at-home features. This is further highlighted by the ROC curves and AUCs obtained (Figure 1). With monitoring and laboratory feature sets, ANN and RF both had AUCs of essentially 1. With the at-home feature set RF perfromed slightly better than ANN with a higher AUC, and a better ROC with a lower variance. Similarly, for creatinine regression both ANN and RF have comparable results using the monitoring and laboratory features, and RF performed better using at-home features.

Exploiting the nature of RF algorithms we extracted the most important features in the classification and regression (Figure 2). Hypertension and diabetes mellitus were the two most important features of the at-home set for both classification and regression, as measured by the mean entropy importance. We had less agreement of the most impactful features between the classification and regression on the monitoring and laboratory feature sets. Hemoglobin was the most important feature for both sets in the classification task, and both red blood cell and hypertension were in the top five for both sets. When predicting creatinine, blood urea was by far the most important feature within the monitoring and laboratory sets, with hemoglobin, red blood cell count, and sodium being in the top five for both sets.

In our study, we leverage a dataset that has been examined in prior works [18, 24–26]. The common thread among these studies, like one of our own, is the focus on CKD classification using all available features. Across these studies and our own (with monitoring or laboratory features), we consistently observe near-perfect accuracy in CKD classification, highlighting the robustness of machine learning methods. Presently, CKD diagnoses require laboratory measurements at least three months apart [6], hence, machine learning could reduce wait time for a diagnosis and treatment plan. Both Khalid *et al*. [25] and Almansour *et al*. [24] explore subsets of this dataset by either using only numerical features or examining the performance with a reduced number of features, respectively. However, our study contributes a novel perspective by categorizing features into at-home, monitoring, and laboratory subsets. Our breakdown sheds light on context-specific importance of attributes. Our approach enhances the interpretability of our models and provides insights into the relevance of features for different aspects of CKD prediction. Qin *et al*. [18] highlight specific gravity, hemoglobin, serum creatinine, albumin, packed cell volume, red blood cell count, hypertension, and diabetes mellitus as key contributors. Our findings overlap significantly with theirs, further validating the importance of these attributes in CKD prognosis and diagnosis. A comprehensive overview of machine learning techniques for CKD classification can be found in [11], where a wide range of methods are assessed. Notably, the best-performing models in their tabulation achieve an accuracy of 98% or higher. Sanmarchi *et al*. [12] conducted a review encompassing 68 relevant articles on CKD prediction, diagnosis, and treatment using a wide range of machine learning methods. Their findings emphasize the importance of attributes such as blood pressure, hemoglobin, sodium, albumin, pus cell, red blood cell count, and diabetes mellitus for CKD prognosis and diagnosis. These highlighted features align with our own observations, underscoring their relevance in the context of CKD assessment.

We employed a similar approach to Wang *et al*. [14] by predicting creatinine levels directly. Their ensemble method achieved an R2 score of 0.5590 using a different dataset, and they emphasize the significance of hemoglobin. In our findings, hemoglobin was in the top three features for both monitoring and laboratory features for creatinine prediction. We note the data used in [14] did not contain blood urea, which we found to be the most significant predictor.

In summary, our study builds upon a dataset examined in previous research and offers a unique perspective by categorizing features into context dependent subsets. Our findings, including the importance of specific attributes and the success of machine learning methods in CKD classification, corroborate and extend upon existing literature, contributing to a better understanding of CKD detection and prediction.

Our study has some limitations however, that warrant consideration. Firstly, while the CKD and not CKD labels in the dataset were assigned by nephrologists using patient history, symptoms, and blood and urine tests, it is not clear precisely what criteria were used to determine the labels. Moreover, the absence of stage-specific information for patients with CKD in our dataset poses a challenge. While we frame creatinine levels as a proxy for CKD stages, this indirect approach may introduce uncertainty, as the correlation between creatinine and estimated GFR may not precisely reflect the true stage of the disease. Additionally, our model’s ability to detect early-stage CKD may be limited, as we primarily focused on classifying CKD in general, not the stage. Having labelled data with explicit CKD stages would enable a more nuanced analysis and classification of disease progression. Presently, our creatinine regression results provide a proxy for stage, but this transition from creatinine to stage introduces additional uncertainty, given the equations involved are empirical in nature. These limitations highlight the need for more comprehensive and stage-specific datasets to further improve the accuracy and clinical relevance of CKD detection and classification models.

## 5 Conclusions

In conclusion, our study represents a step toward leveraging machine learning for the early detection and classification of CKD, addressing a pressing concern in clinical medicine. By examining different feature subsets of at-home, monitoring, and laboratory, we offer insights into the potential use of such models in diverse clinical settings. Our findings, which align with previous research, stress the importance of specific features such as blood urea, hemoglobin, blood pressure, and diabetes mellitus in CKD detection and classification. Looking ahead, the impact of this work extends to the development of more robust and accurate CKD screening tools, potentially facilitating earlier interventions and improved patient outcomes. However, we recognize the need for larger and more comprehensive datasets, including detailed CKD stage information, to enhance the precision of our models. Additional, high-quality data will help improve the accuracy of ANN CKD classification accuracy. Furthermore, training a dual-task ANN to simultaneously classify CKD and predict creatinine may improve the performance of both tasks. With continued research and access to richer clinical data the integration of machine learning techniques into routine CKD diagnosis and prognosis holds the promise of assisting the field of nephrology. Machine learning could improve the lives of many individuals affected by this pervasive health condition.

## A Descriptive statistics of dataset

See Table 5 for descriptive statistics of the numerical features and Table 6 for descriptive statistics of the nominal features.

**Table 5:** Descriptive statistics of numerical features.

|  | age | bp | bgr | bu | sc | sod | pot | hemo | pcv | wbcc | rbcc |
| --- | --- | --- | --- | --- | --- | --- | --- | --- | --- | --- | --- |
| count | 391 | 388 | 356 | 381 | 383 | 313 | 312 | 348 | 329 | 294 | 269 |
| mean | 51.48 | 76.47 | 148.04 | 57.43 | 3.07 | 137.53 | 4.63 | 12.53 | 38.88 | 8406.12 | 4.71 |
| std | 17.17 | 13.68 | 79.28 | 50.50 | 5.74 | 10.41 | 3.19 | 2.91 | 8.99 | 2944.47 | 1.02 |
| min | 2.00 | 50.00 | 22.00 | 1.50 | 0.40 | 4.50 | 2.50 | 3.10 | 9.00 | 2200.00 | 2.10 |
| 25% | 42.00 | 70.00 | 99.00 | 27.00 | 0.90 | 135.00 | 3.80 | 10.30 | 32.00 | 6500.00 | 3.90 |
| 50% | 55.00 | 80.00 | 121.00 | 42.00 | 1.30 | 138.00 | 4.40 | 12.65 | 40.00 | 8000.00 | 4.80 |
| 75% | 64.50 | 80.00 | 163.00 | 66.00 | 2.80 | 142.00 | 4.90 | 15.00 | 45.00 | 9800.00 | 5.40 |
| max | 90.00 | 180.00 | 490.00 | 391.00 | 76.00 | 163.00 | 47.00 | 17.80 | 54.00 | 26400.00 | 8.00 |

**Table 6:**
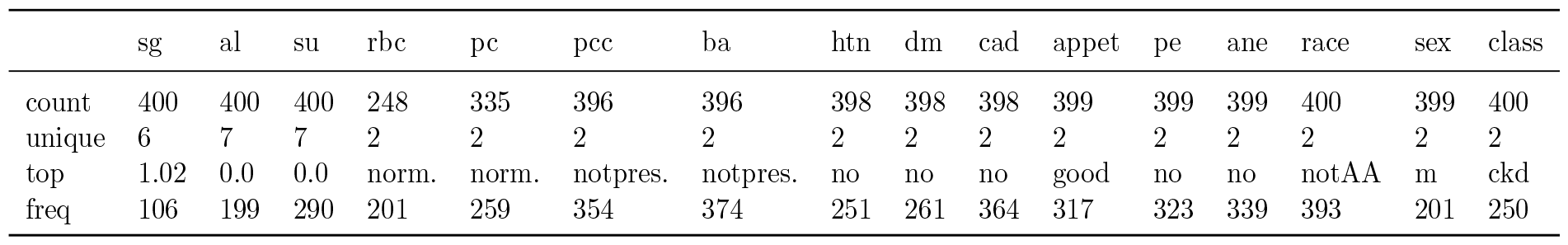
Descriptive statistics of nominal features.

## Data Availability

All data produced in the present study are available upon reasonable request to the authors

## List of abbreviations

ANN: Artificial neural network
AUC: Area under curve
CKD: Chronic kidney disease
FNR: False negative rate
FPR: False positive rate
GFR: Glomerular filtration rate
*k*-NN: *k*-nearest neighbours
MAE: Mean absolute error
MSE: Mean squared error
R2: R-squared
RF: Random forest
ROC: Receiver operator curves
TNR: True negative rate
TPR: True positive rate

## Acknowledgements

The authors acknowledge the support provided by the EPSRC Centre for Doctoral Training in Industrially Focused Mathematical Modelling (EP/L015803/1). The authors would like to thank Markus Dablander for useful discussions and critical reading of the manuscript. The authors would like to thank Vironix and the participants at the 39th Annual Mathematical Problems in Industry (MPI) workshop, where this problem was first presented.

## Author contributions

BM: data processing, ANN model training, visualizations. AKB: RF model training, RF feature importance. GSB: RF model training. All authors designed the studies, interpreted results, and drafted and reviewed the manuscript.

## References

[1] Grams, M.E., Chow, E.K.H., Segev, D.L., Coresh, J.: Lifetime incidence of CKD stages 3–5 in the United States. American Journal of Kidney Diseases 62(2), 245–252 (2013). 10.1053/j.ajkd.2013.03.009

[2] Vassalotti, J.A., Stevens, L.A., Levey, A.S.: Testing for chronic kidney disease: A position statement from the National Kidney Foundation. American Journal of Kidney Diseases 50(2), 169–180 (2007). 10.1053/j.ajkd.2007.06.013

[3] Qezelbash-Chamak, J., Badamchizadeh, S., Eshghi, K., Asadi, Y.: A survey of machine learning in kidney disease diagnosis. Machine Learning with Applications 10 (2022). 10.1016/j.mlwa.2022.100418

[4] Hogg, R.J., Furth, S., Lemley, K.V., Portman, R., Schwartz, G.J., Coresh, J., Balk, E., Lau, J., Levin, A., Kausz, A.T., Eknoyan, G., Levey, A.S.: National Kidney Foundation’s kidney disease outcomes quality initiative clinical practice guidelines for chronic kidney disease in children and adolescents: Evaluation, classification, and stratification. Pediatrics 111(6), 1416–1421 (2003). 10.1542/peds.111.6.1416

[5] Inker, L.A., Astor, B.C., Fox, C.H., Isakova, T., Lash, J.P., Peralta, C.A., Tamura, M.K., Feldman, H.I.: KDOQI US commentary on 12 ML for classifying CKD and predicting creatinine the 2012 KDIGO clinical practice guideline for the evaluation and management of CKD. American Journal of Kidney Diseases 63(5), 713–735 (2014). 10.1053/j.ajkd.2014.01.416

[6] Levey, A.S., Coresh, J., Balk, E., Kausz, A.T., Levin, A., Steffes, M.W., Hogg, R.J., Perrone, R.D., Lau, J., Eknoyan, G.: National Kidney Foundation practice and guidelines for chronic and kidney and disease: Evaluation and classification and stratification. Annals of Internal Medicine 139(2), 137–147 (2003). 10.7326/0003-4819-139-2-200307150-00013

[7] Levey, A.S., Bosch, J.P., Lewis, J.B., Greene, T., Rogers, N., Roth, D.: A more accurate method to estimate glomerular filtration rate from serum creatinine: A new prediction equation. Annals of Internal Medicine 130(6), 461–470 (1999). 10.7326/0003-4819-130-6-199903160-00002

[8] Levey, A.S., Stevens, L.A., Schmid, C.H., Zhang, Y.L., III, A.F.C., Feldman H.I., Kusek, J.W., Eggers, P., Lente, F.V., Greene, T., Coresh, J., for the CKD-EPI (Chronic Kidney Disease Epidemiology Collaboration): A new equation to estimate glomerular filtration rate. Annals of Internal Medicine 150(9), 604–612 (2009). 10.7326/0003-4819-150-9-200905050-00006

[9] Deo, R.C.: Machine learning in medicine. Circulation 132(20), 1920–1930 (2015). 10.1161/CIRCULATIONAHA.115.001593

[10] Badrouchi, S., Bacha, M.M., Hedri, H., Ben Abdallah, T., Abderrahim, E.: Toward generalizing the use of artificial intelligence in nephrology and kidney transplantation. Journal of Nephrology 36(4), 1087–1100 (2023). 10.1007/s40620-022-01529-0

[11] Nimmagadda, S.M., Agasthi, S.S., Shai, A., Khandavalli, D.K.R., Vatti, J.R.: Kidney failure detection and predictive analytics for ckd using machine learning procedures. Archives of Computational Methods in Engineering 30(4), 2341–2354 (2022). 10.1007/s11831-022-09866-w

[12] Sanmarchi, F., Fanconi, C., Golinelli, D., Gori, D., Hernandez-Boussard, T., Capodici, A.: Predict, diagnose, and treat chronic kidney disease with machine learning: A systematic literature review. Journal of Nephrology 36, 1101–1117 (2023). 10.1007/s40620-023-01573-4

[13] Schena, F.P., Anelli, V.W., Abbrescia, D.I., Di Noia, T.: Prediction of chronic kidney disease and its progression by artificial intelligence algorithms. Journal of Nephrology 35(8), 1953–1971 (2022). 10.1007/s40620-022-01302-3

[14] Wang, W., Chakraborty, G., Chakraborty, B.: Predicting the risk of chronic kidney disease (CKD) using machine learning algorithm. Applied Sciences 11(1), 202 (2020). 10.3390/app11010202

[15] Rubini, L., Soundarapandian, P., Eswaran, P.: Chronic Kidney Disease. UCI Machine Learning Repository (2015). 10.24432/C5G020

[16] Rubini, L.J.: An optimal feature selection and intelligent classification methods for chronic kidney disease prediction. PhD thesis, Alagappa University (2021). http://hdl.handle.net/10603/391737

[17] Ilyas, H., Ali, S., Ponum, M., Hasan, O., Mahmood, M.T., Iftikhar, M., Malik, M.H.: Chronic kidney disease diagnosis using decision tree algorithms. BMC Nephrology 22(1) (2021). 10.1186/s12882-021-02474-z

[18] Qin, J., Chen, L., Liu, Y., Liu, C., Feng, C., Chen, B.: A machine learning methodology for diagnosing chronic kidney disease. IEEE Access 8, 20991–21002 (2020). 10.1109/access.2019.2963053

[19] Fawcett, T.: An introduction to ROC analysis. Pattern Recognition Letters 27(8), 861–874 (2006). 10.1016/j.patrec.2005.10.010

[20] Hogan, J., Adams, N.M.: On averaging ROC curves. Transactions on Machine Learning Research (2023)

[21] Chollet, F., et al.: Keras (2015). https://keras.io

[22] Pedregosa, F., Varoquaux, G., Gramfort, A., Michel, V., Thirion, B., Grisel, O., Blondel, M., Prettenhofer, P., Weiss, R., Dubourg, V., Vanderplas, J., Passos, A., Cournapeau, D., Brucher, M., Perrot, M., Duchesnay, E.: Scikit-learn: Machine learning in Python. Journal of Machine Learning Research 12, 2825–2830 (2011)

[23] O’Malley, T., Bursztein, E., Long, J., Chollet, F., Jin, H., Invernizzi, L., et al.: Keras-Tuner. GitHub (2019). https://github.com/keras-team/keras-tuner

[24] Almansour, N.A., Syed, H.F., Khayat, N.R., Altheeb, R.K., Juri, R.E., Alhiyafi, J., Alrashed, S., Olatunji, S.O.: Neural network and support vector machine for the prediction of chronic kidney disease: A comparative study. Computers in Biology and Medicine 109, 101–111 (2019). 10.1016/j.compbiomed.2019.04.017

[25] Khalid, H., Khan, A., Khan, M.Z., Mehmood, G., Qureshi, M.S.: Machine learning hybrid model for the prediction of chronic kidney disease. Computational Intelligence and Neuroscience 2023, 1–14 (2023). 10.1155/2023/9266889

[26] Pal, S.: Prediction for chronic kidney disease by categorical and non categorical attributes using different machine learning algorithms. Multimedia Tools and Applications (2023). 10.1007/s11042-023-15188-1

